# Genomic investigation of MRSA bacteremia relapse reveals diverse genomic profiles but convergence in bacteremia-associated genes

**DOI:** 10.1101/2025.03.24.25324140

**Authors:** Brooke M. Talbot, Natasia F. Jacko, Katrina S. Hofstetter, Tara Alahakoon, Kevin Bouiller, Timothy D. Read, Michael Z. David

**Author notes:** Correspondance should be directed to Michael Z. David, M.D., Ph.D., Division of Infectious Diseases, Department of Medicine, University of Pennsylvania 423 Guardian Drive, Room 707, Philadelphia, PA, USA 19104. Alternate Contact: Timothy D. Read, 1750 Haygood Dr., Atlanta, GA, USA 30322.

## Abstract

**Background:** Recurrence of methicillin-resistant *Staphylococcus aureus* (MRSA) bacteremia is a high risk complication for patients. Distinguishing persistent lineages from new infections is not standardized across clinical studies.

**Methods:** We investigated factors contributing to recurrence of MRSA bacteremia among subjects in Philadelphia, Pennsylvania. Subject demographics and clinical history were collected and paired with whole-genome sequences of infection isolates. Recurrent bacteremia episodes were recorded and defined as relapse infections (same lineage) or new infections by genomic criteria, where a relapse contains isolates <=25 single nucleotide polymorphisms (SNP) different, and by clinical criteria. All isolates were assessed for pairwise SNP distances, common mutations, and signatures of within-host adaptation using the McDonald-Kreitman test. Clusters of transmission between relapse-associated isolates and other subject lineages were identified.

**Results:** Among 411 sequential subjects with MRSA bacteremia, 32 experienced recurrent bacteremia episodes, with 24 subjects having exclusively relapse infections, six with infections exclusively from a new strain, and two patients with both relapse and new infections. No concordance between a genomic and a clinical definition of relapse was evident (Cohen’s Kappa = 0.18, CI: -0.41). Recurrence-associated lineages exhibited signatures of positive selection(G-test:<0.01) . Genes with SNPs occurring in multiple relapse lineages have roles in antibiotic resistance and virulence, including 5 lineages with mutations in *mprF* and 3 lineages with mutations in *rpoB,* which corresponded with evolved phenotypic changes in daptomycin and rifampin resistance.

**Conclusions:** Recurrent infections have a diverse strain background. Relapses can be readily distinguished from newly acquired infections using genomic sequencing but not clinical criteria.

## Introduction

*Staphylococcus aureus* bacteremia is a complex clinical syndrome which often leads to adverse patient outcomes, including endocarditis and other metastatic infections [1]. Recurrence of *S. aureus* bacteremia, when patients are diagnosed with bacteremia again after assumed resolution of a previous infection, is an ongoing clinical challenge. Global data demonstrate that 5 - 15 percent methicillin-resistant *S. aureus* (MRSA) bacteremia (MRSAB) episodes result in recurrence, and the risks associated with recurrence are similarly heterogenous to bloodstream infections overall [2–9]. Known risk factors for recurrence of *S. aureus* bacteremia in adults include younger patient age, presence of a foreign body, hemodialysis dependence, valvular heart disease, liver cirrhosis, and endocarditis [6,9].

Distinguishing new infections after clearance from cryptically persisting bacteria within host tissues is a challenge for clinicians, especially if the focus of an infection is unknown. We split recurrent MRSAB into two types: “relapses”, caused by the same genetic lineage, and “new infections” in which a different *S. aureus* lineage is responsible. Recurrence has been heterogeneously defined using strain genotypes (e.g., *spa* typing or multilocus sequence typing [MLST]), time intervals, or clinical history of previous infections [4,6–9]. Without including genomic similarity criteria, it has not been possible to distinguish relapse from new infections accurately. Consequently, misidentification of the two types of recurrence includes, for relapses, not identifying persistent infections, and for new infections, that a patient may have a new risk factor for *S. aureus* infection. Defining the nature of a recurrence may help therapeutic approaches for preventing subsequent infections.

Colonization by *S. aureus* has been associated with higher risk of developing bacteremia. The transition from colonization to invasive infection, and the subsequent treatment regimen, can result in bacterial adaptive evolution [10–12]. Mutations have been found to have occurred convergently in multiple different MRSAB isolates. These genetic changes alter virulence regulation [13–25] and antibiotic targets [13–15,26,27] and result in the development of small colony variants [28,29] among other phenotypes. Emerging mutations can confer cross resistance to first-line antibiotic treatments or multidrug resistance, further complicating prevention of persistent infections or cure in patients with deep-seated foci [10]. Several loci appear to increase the risk for metastatic and persistent infections, and possible recurrence. An array of putative loci have been identified to adapt during *S. aureus* bacteremia, but it is not known which, if any, are more associated with relapse versus new infection. There is also little information on how *S. aureus* strain background influences forward evolution during bacteremia or the propensity for relapse. Knowledge of genetic change patterns in *S. aureus* during bacteremia episodes may help differentiate new infections from relapses.

To define genetic differences and risks for MRSAB recurrence, we examined a cohort of patients in Philadelphia, Pennsylvania experiencing MRSAB during a five-year period who received care at a single hospital system. We performed whole-genome sequencing (WGS) on single isolates from single MRSAB episodes and recurrent episodes. We examined the associations between host clinical factors and bacterial genetics to characterize recurrent MRSAB, and defined the transmission and adaptive history of infections that resulted in a relapse.

## Methods

### Subject cohort and isolate collection

This study was considered exempt by the University of Pennsylvania Institutional Review Board. Subject MRSAB isolates were included from a cohort of adult patients admitted to at least one of two hospitals in Philadelphia, Pennsylvania and diagnosed with MRSAB between July 2018 and February 2022. A single colony representative isolate of the first positive culture was stored for each MRSAB episode. Clinical and demographic information was collected through medical record reviews for all infections, including age at time of isolate collection, race, ethnicity, sex at birth, death within 30 days of infection diagnosis, comorbidities, complications from bacteremia, antibiotic treatment, suspected source site of infection, and healthcare-associated epidemiological category. Antibiotic resistance phenotypes were collected from clinical microbiology records associated with the unique MRSA isolates.

Antibiotic resistance was assessed using the Vitek 2 automated system, and assigned susceptibility/resistance in accordance with Clinical and Laboratory Standards Institute protocols [30].

### Clinical distinction of relapse and new infections

Subsequent MRSAB events per subject (a “recurrent episode”) were classified as a “relapse” or “new infection” according to the following criteria as outlined in Figure S1: MRSAB episodes include all isolates collected in a 30 day period from the first episode isolate. At each discrete medical encounter during which MRSAB was detected, subjects with one or more MRSA isolate from blood were assessed for previous MRSAB and subdivided into discrete bacteremia episodes if greater than 30 days apart. Subjects with multiple discrete bacteremia episodes were considered to have one or more “recurrent episodes.” Recurrent episodes were then categorized into “new infections” and “relapse” infections using clinical criteria or genomic criteria. For clinical criteria, new infections had to fulfill all of the following: The first positive culture from the episode was 30 days or more following the last positive MRSA blood culture, all symptoms at the source site and metastatic sites of the previous infection were resolved, no new antibiotics were prescribed after completion of definitive therapy for the index MRSAB episode aside from antibiotics aimed at suppression of an infected focus, if the subject was administered suppressive antibiotics the infection site (source) was different from the previous infection, if a central venous catheter was exchanged at the time of the previous MRSAB then it was not exchanged over a wire but instead was resited [31], or the infection source was clinically ruled to be different from the previous infection. Otherwise, if any one or more of these criteria were not true the bacteremia episode was considered a relapse.

### Sequencing quality and phylogenetics

Genomic DNA was extracted from *S. aureus* isolates and sequenced at the Penn/Children’s Hospital of Philadelphia Microbiome Center, using paired-end short read whole-genome sequencing as previously described [32]. Reads were processed using Bactopia (v 3.0.0) [33]. Briefly, adaptor sequences were removed, and reads were de novo assembled using Shovill (v.1.1.0). Sequences were used for further investigation if reads had at least an average per-read quality score of Q12 and a mean read length of 49bp, and the genomic assembly had at least 20x coverage and no more than 500 contigs. Multilocus sequence type (ST) and clonal complex (CC) were assigned by calling Ariba (v 2.14.6) within Bactopia, which utilized the *S. aureus* specific ST scheme from PubMLST [34] . For novel STs or STs without a defined CC, the CC was manually defined using a maximum likelihood (ML) tree and relative position to the next isolate with a defined CC. A core genome alignment was created among all MRSAB isolates and MSSA Strain Newman (GCF_020985245.1) as an outgroup with the Bactopia subworkflow “pangenome” using PIRATE [35]. Areas of likely recombination were identified and masked using ClonalFrame ML (v.1.12) [36]. A ML tree of all isolates was created from the masked alignment with IQTree (2.1.2) [37] and ModelFinder [38], which determined the best fit model to be a generalized time reversible model with Empirical base frequencies plus the FreeRate model.

### Genomic criteria for relapsed and new infections and cluster identification

Snp-dists (0.8.2) was used to calculate single nucleotide polymorphism (SNP) pairwise distances between aligned isolates. Subjects with recurrent MRSAB episodes were categorized as having a relapsed or new infection based on pairwise SNP differences from the isolate of the most recent previous episode relative to the next subsequent episode. A conservative SNP threshold of <=25 SNPs between episodes defined a relapse based on previous observations of SNP distances of MRSA lineages persisting within a host [39]. The phylogenetic branch patterns were also investigated between recurrent episode isolates: relapse isolates must also share their most-recent common ancestor with another isolate from within the same human subject and share the same ST. Cohen’s kappa [40,41] was calculated between the clinical and genomic definitions of relapse to assess method agreement and clinical criteria sensitivity and specificity relative to the genomic criteria.

### Association of clusters with demographic and clinical data

Demographic and clinical characteristics were compared between subjects with recurrent MRSAB episodes and single MRSAB episodes using Kruskal-Wallis test for continuous variables and Peason’s Chi-square test or Fisher’s exact test for categorical variables.

Demographic and clinical history were first compared between MRSAB subjects who died within their index MRSAB episode window (30 days) and those who survived their index infection (Table S1). Decedents from index MRSAB infection were excluded from the analysis assessing risk factors associated with MRSAB infection survival.

Genomic similarity between subjects’ isolates was also compared to identify potential transmission clusters. For all relapse-associated isolates <= 25 SNPs from different subjects’ isolates, subtrees of the larger ML tree were created and examined for branch structure and days since the earliest collection date in the cluster to identify the role of relapse isolates in putative onward transmission.

### Variant calling in relapse lineages

Bactopia’s Snippy subworkflow, which utilizes Snippy v4.6.0 [42], was used for variant calling of SNPs of all isolates against a previously generated ancestrally reconstructed *S. aureus* genome [43]. Variant calling was also conducted for each set of identified relapse infections by using the first known isolate (index) in each lineage as the reference. Index isolates were annotated using Bakta (v1.9.3)[44] to generate the reference sequence for variant analysis. The – snippy-core option was run on all isolates to identify variants in core coding regions across all genomes to produce an alignment and predicted changes to amino acid structure. To identify commonly occurring non-synonymous changes within each recurrence lineage, the –snippy-core option was also used to create a SNP alignment for each set of isolates from the same subject to identify common variants and changes to amino acid structure. SNPs in coding regions were concatenated and summarized for all lineages.

### Creating a database of bacteremia-associated mutations

A literature review was conducted to create a database of previously identified genes and/or mutations in *S. aureus* genomes associated with bacteremia. PubMed was searched in December 2023 using the query (((staphylococcus aureus)) AND ((bacteremia) OR (bloodstream infection))) AND (genetic mutation). Only peer-reviewed articles were scanned for evidence. Genes or mutations in genes were considered if the study reported that they occurred in the *S. aureus* genome, samples were derived from *S. aureus* bacteremia, were associated with characteristics relevant to an outcome (e.g., virulence expression or host survival) or were associated with host phenotypic changes related. A listing of citations identifying these genes is in Table S2.

### Calculation of the index of neutrality

McDonald-Kreitman tests [45] were conducted on all MRSAB isolates and the subset of relapse-associated isolates to assess selection on the whole genome or on known bacteremia-associated genes. A core genome alignment was created using snippy-core for all isolates and for isolates associated with a relapse lineage. The ancestral reference served as the outgroup. Fixed sites were determined by counting the nucleotide sites universally conserved from the reference among compared isolates. Polymorphic sites were determined if there were nucleotide sites that differed between individuals within the isolate groups. A G-test was used to evaluate the significance of the neutrality index.

## Results

### Recurrent MRSAB are from common clinical and phylogenetic backgrounds among circulating strains

We sequenced 456 *S. aureus* isolates from episodes of MRSAB and included 411 subjects, of whom 32 had at least 1 recurrent MRSAB episode in addition to the index infection (77 isolates). Recurrence-associated isolates occurred across major CCs of the *S. aureus* species, including CC5 (N = 29), CC8 (N = 41), CC30 (N = 3), CC78 (N = 3) and CC72 (N = 1) (Fig. 1). CC distribution was similar between recurrence and non-recurrence isolates (p=0.91), and there was no difference in the number of recurrences over time.

**Figure 1:**
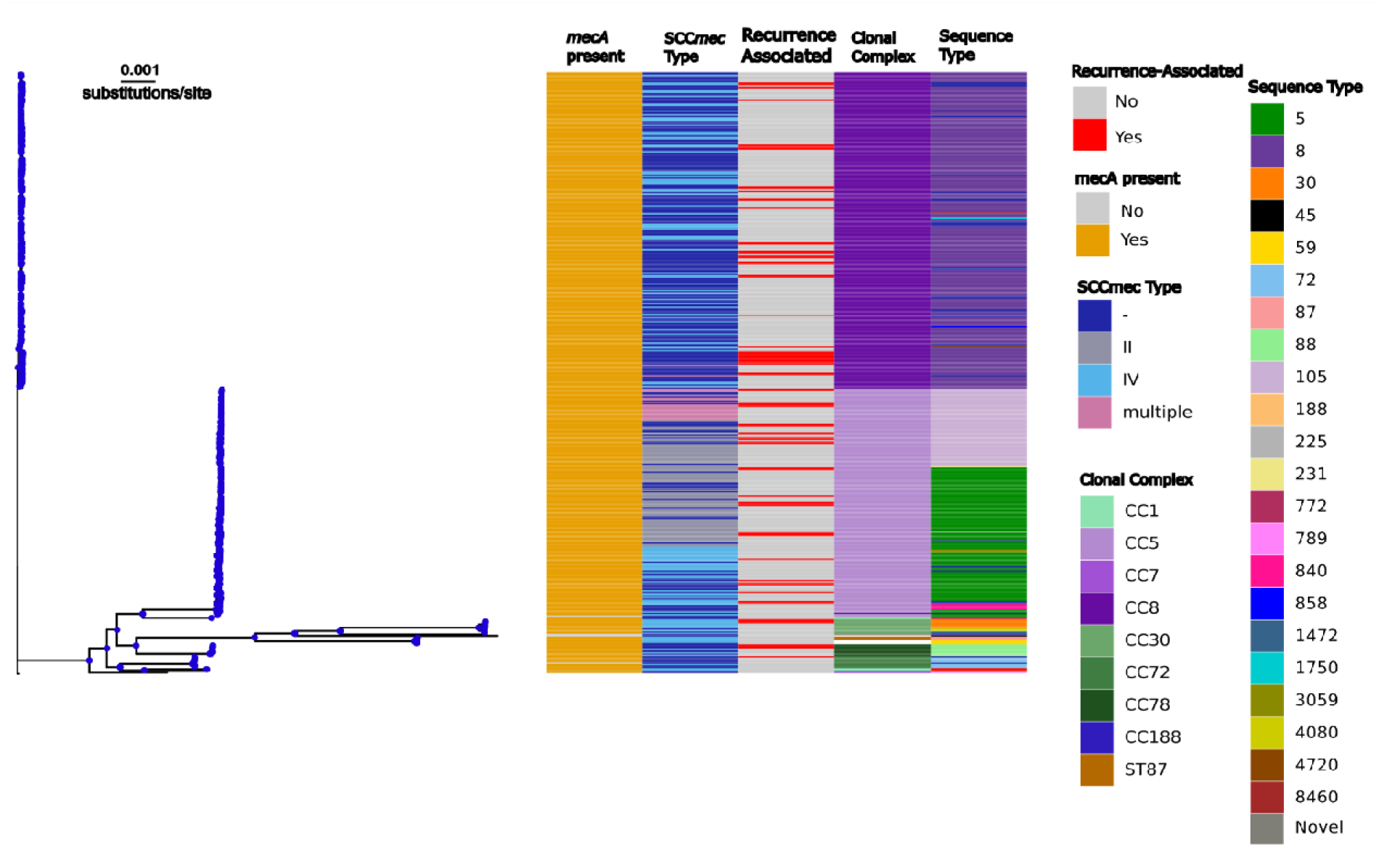
Recurrent MRSAB isolates share a similar phylogenetic distribution to non- recurrent MRSAB episodes. A core pangenome tree of 411 MRSA bloodstream isolates was constructed and rooted using MSSA strain Newman (GCF_020985245.1). Whether or not an isolate was associated with a recurrent episode and molecular characteristics are shown in the heat map including, from left to right, the presence or absence of *mecA*, *mecA* type, clonal complex, and mutlilocus sequence type (ST). Blue dots indicated nodes where Shimodaira- Hasegawa approximate likelihood ratio test and ultrafast bootstrap values were >70).

Subjects surviving the index MRSAB episode (N=311) were assessed for risk factors for recurrent MRSAB (Table 1). When subjects with recurrent episodes were compared to those with no record of recurrence, we found no significant differences in clinical or demographic characteristics. Subjects epidemiologically categorized as having healthcare-acquired community onset (HACO) infections were more frequent among recurrent subjects (84%) compared to non-recurrence patients (66%), although these associations were not statistically significant.

**Table 1.**
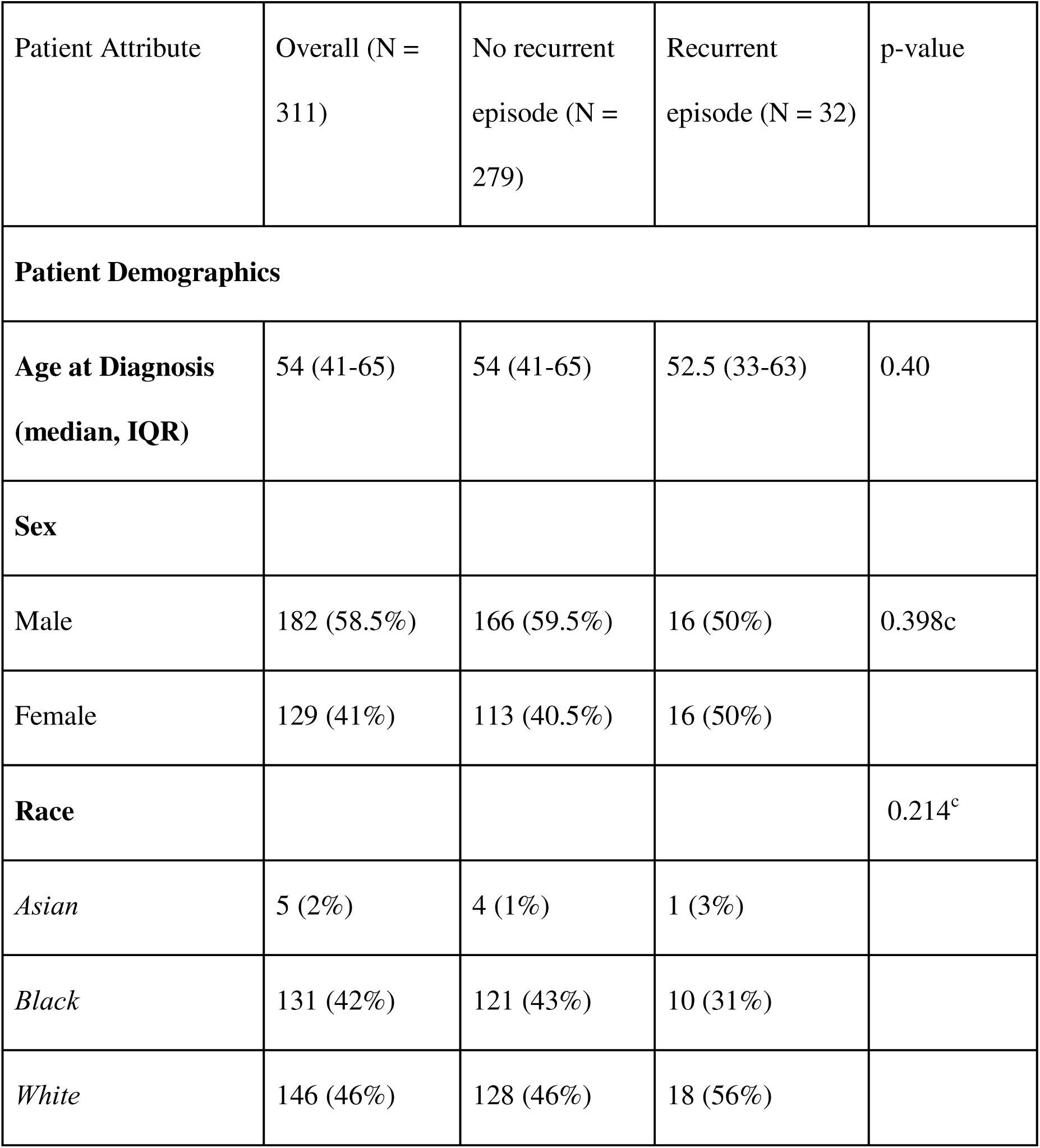
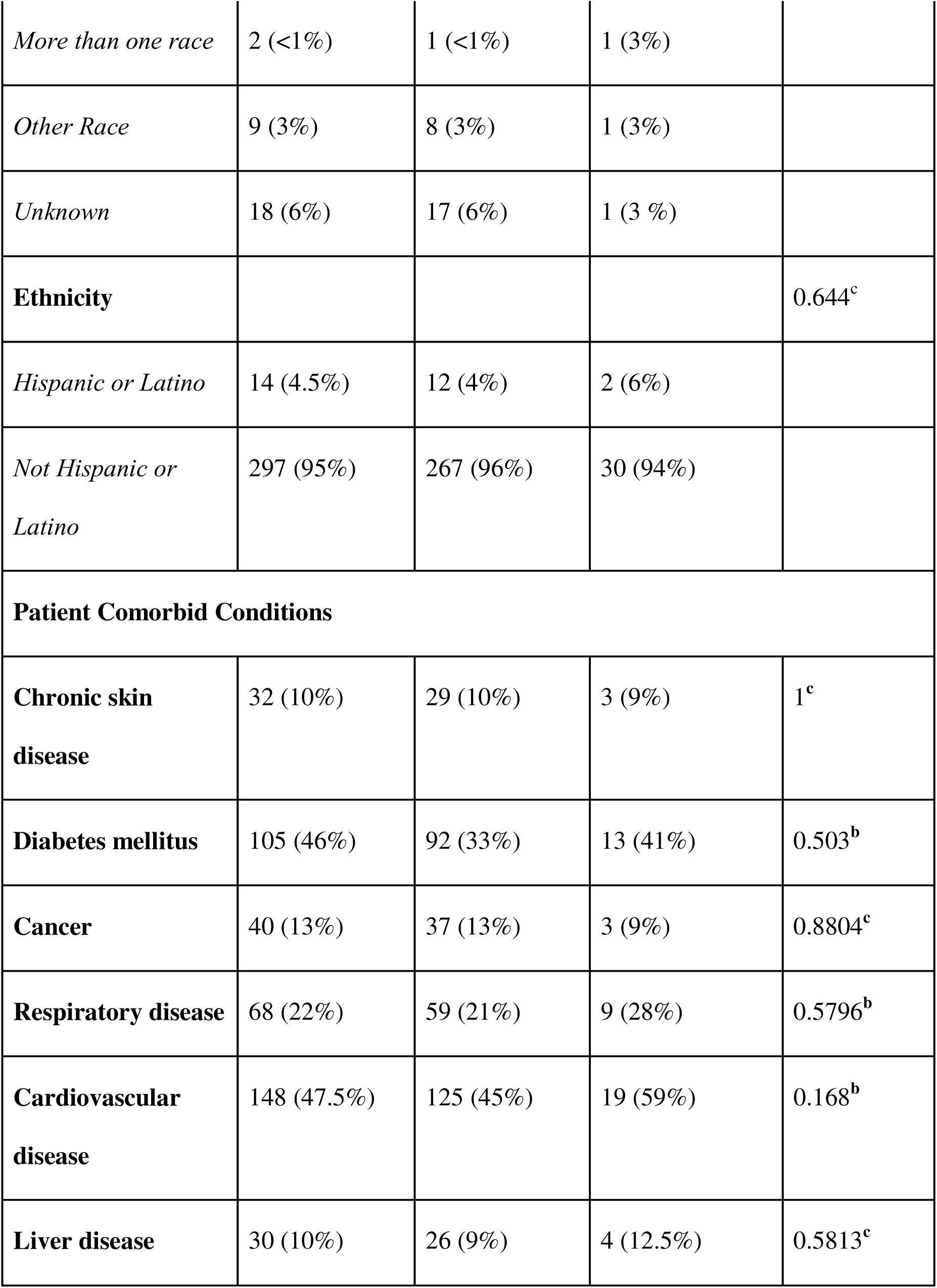
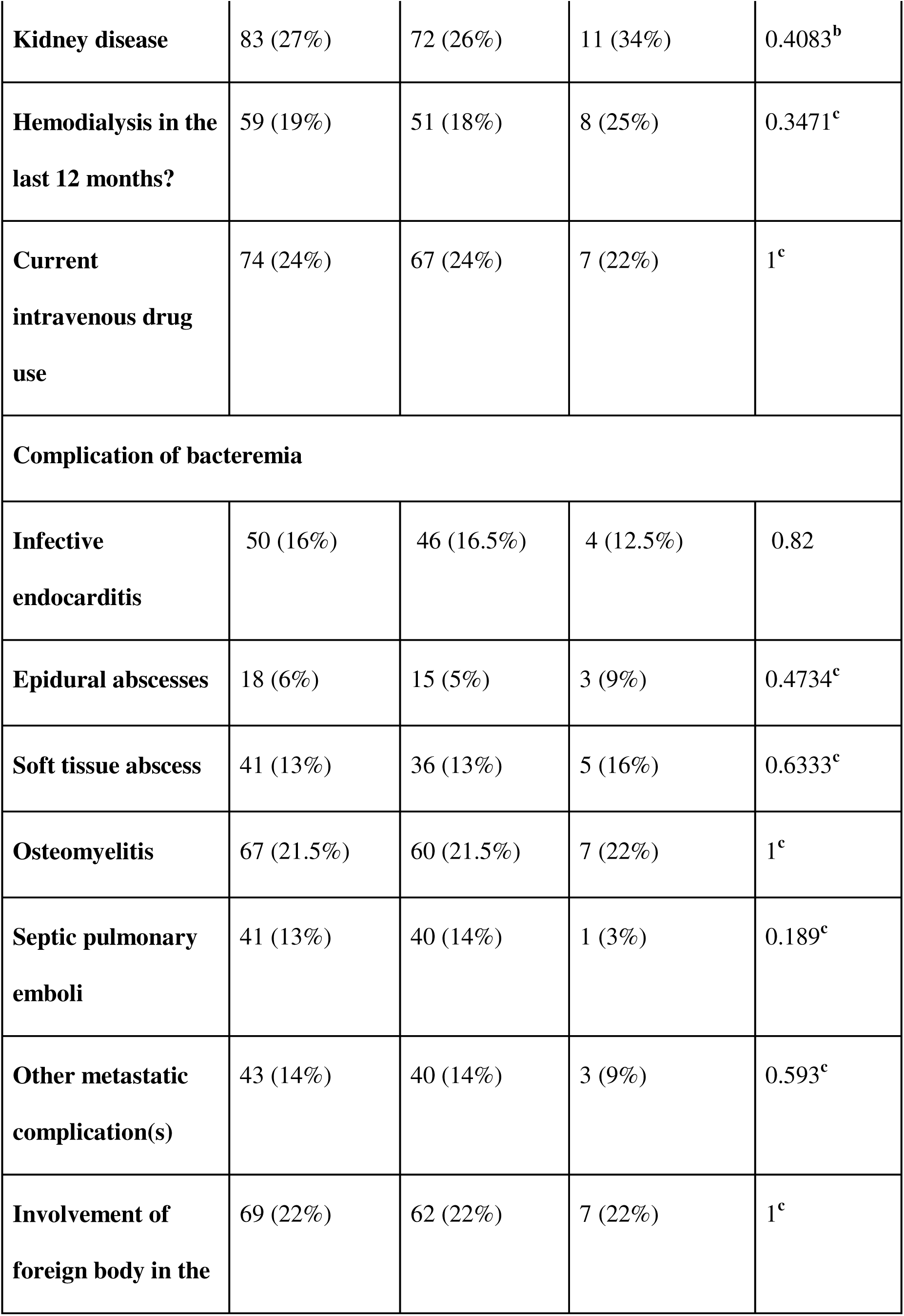
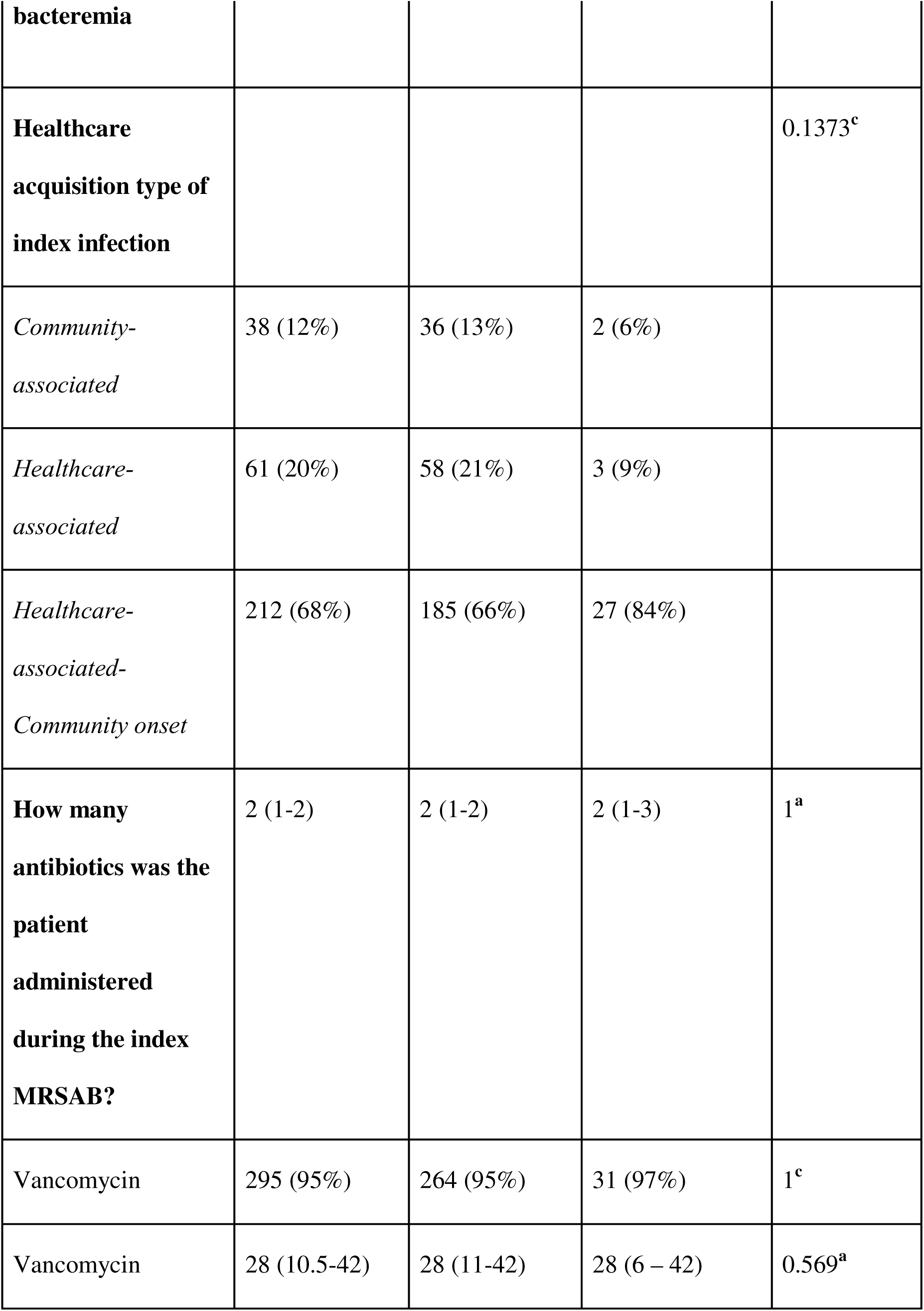
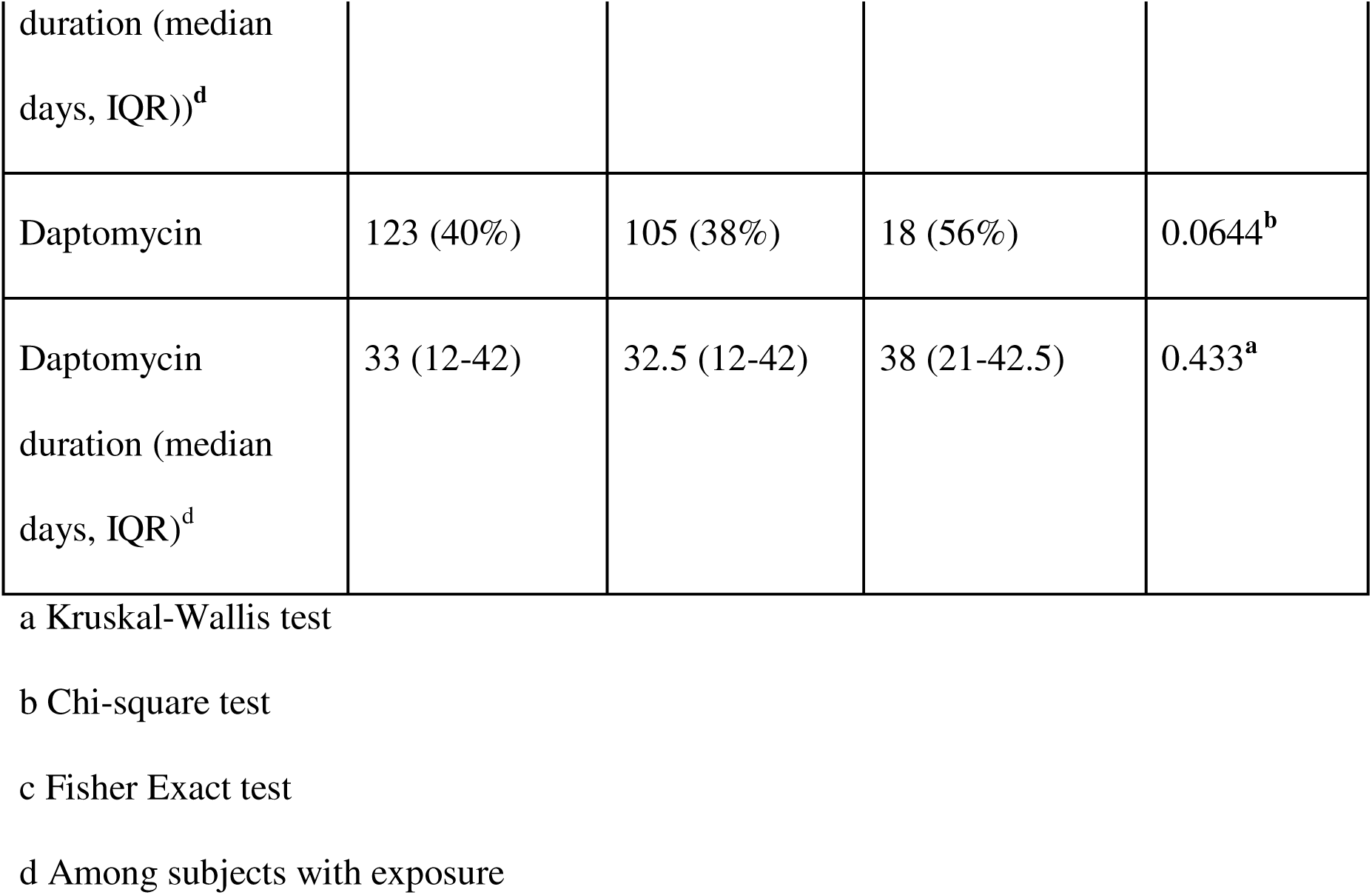
Clinical and demographic characteristics of subjects at the time of the first MRSA bacteremia episode for 311 subjects surviving this episode, comparing those with a recurrent and those without a recurrent MRSA bacteremia episode.

### New infections are genetically distinct from relapse infections among recurrent episodes

Using genomic criteria we classified 24 subjects as having only relapse infections and six with infections only from a new strain; two subjects experienced both relapse and new infections. We identified 45 subsequent recurrent episodes among the 32 subjects. Relapse episodes occurred sooner after the prior infection episode (N = 37, median 97 days, interquartile range, IQR 69-200 days) compared to new strain episodes (N = 8, median 311 days, IQR 79-568 days), though this was not statistically significant (P=0.17) (Fig. 2A). Most relapses fell well below the 25 SNP threshold, with only 3 episodes defined instead by their shared common ancestor with intrasubject isolates. All isolates classified as new infections were distantly related to the subject’s previous infection and frequently belonging to a separate clade of *S. aureus* entirely (Fig. 2B).

**Figure 2:**
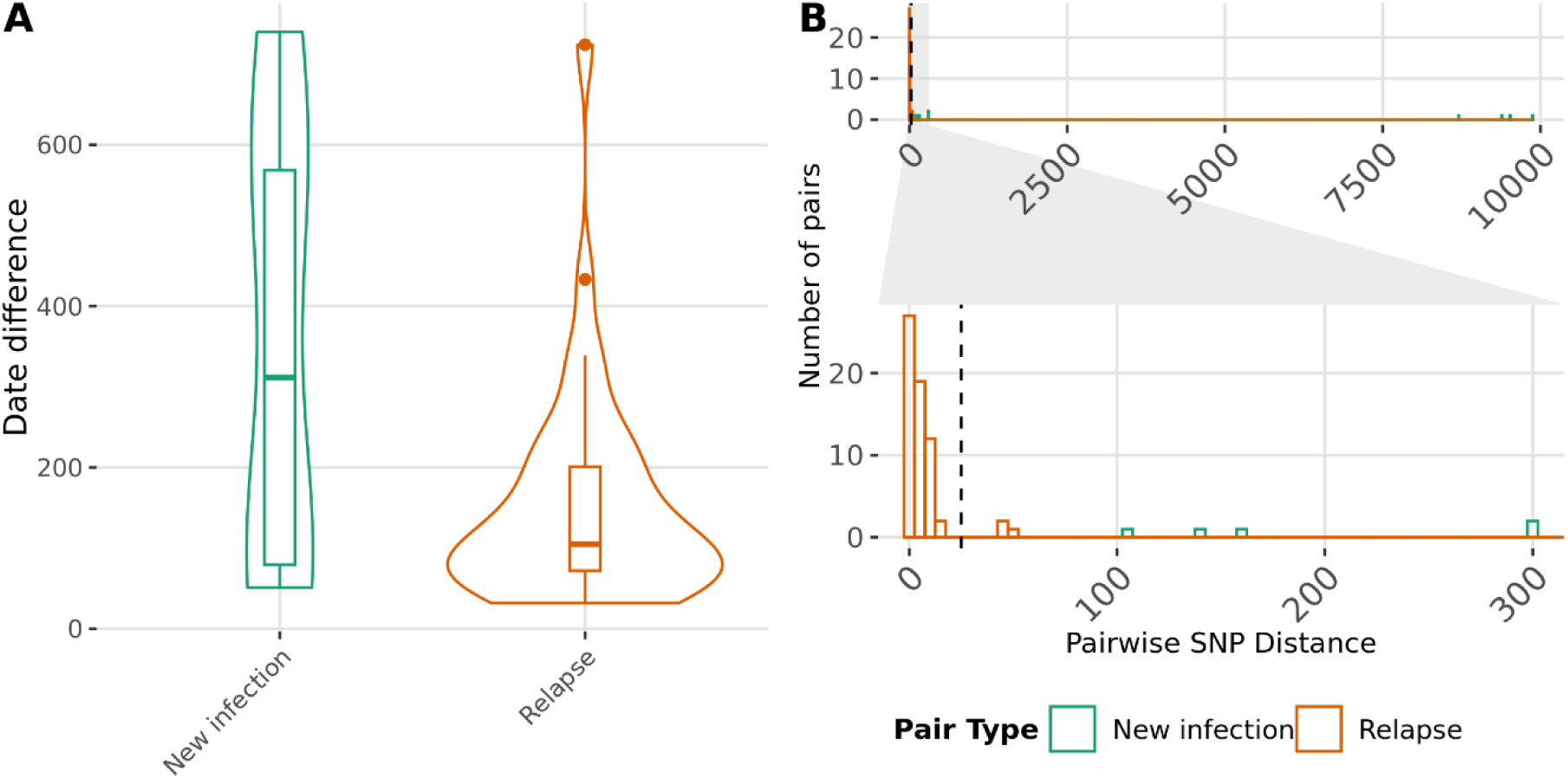
Relapsing infections and new infections within a patient are genomically distinguishable. **Subsequent** isolate pairs among recurrences episodes (n=45) were separated into relapse-associated (n=37) or new infections (n=8) based on pairwise single nucleotide polymorphism (SNP) distance and distance to last common ancestor between isolates within the subject (A) Difference in time between subsequent episodes for genomically new infections and relapse associated infections were compared using the Kruskal-Wallice test (p=0.17). (B) Counts of the pairwise SNP distances between all isolate pairs from the same host were quantified (N=73). The inset display demonstrates counts where the SNP distance between isolates was between 0 and 350 SNPs. The dotted line represents the 25 SNP threshold.

When clinical and genomic definitions were compared, the overall concordance was poor (Cohen’s Kappa = 0.18, CI: -0.41), with the genomic definition predicting that 82% of subsequent infections were related to the previous infection, and the clinical definition predicting that 50% were related (Fig. 3). Using genomics as a gold standard for relapse, the clinical definition had a sensitivity of 55% and a specificity of 75%. When a device or foreign body was implicated in any infection, however, there was often high concordance identifying relapse infections. The genomic definitions of relapse and new infections were used for the remainder of analyses.

**Figure 3:**
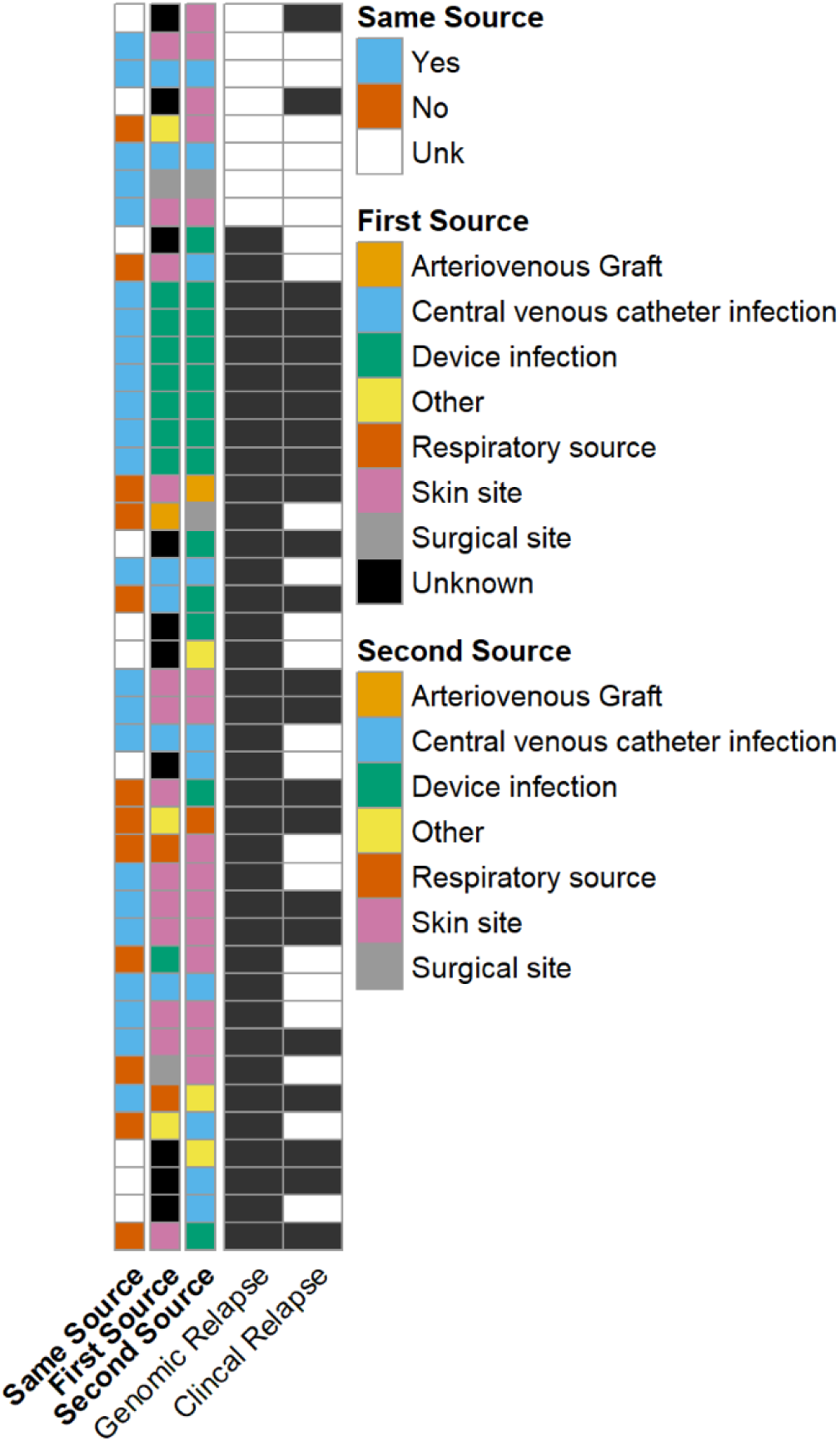
Clinical and genomic definitions of relapse are discordant. Pairs of isolates from all recurrent infections were compared and identified as relapse (filled black rectangle) or new infections (white rectangles) based on a genomic definition or clinical definition, for a total of 45 pairs. The suspected source type of each infection within the episode was identified, with the least recent isolate associated with the “First Source” and the most recent isolate associated with the “Second Source.” Clinical source type was additionally assessed to determine if the first and second source were anatomically from the same source.

### Relapse infections demonstrate distinct adaptation to the host

Across all isolates, there was a trend of neutral evolution across the genome and in the subset of genes associated with bacteremia. Comparatively, relapse-associated lineages showed a significant signature of positive selection in the whole genome (Table 2). Although relatively few bacteremia-associated genes were synonymously mutated, there were notably no synonymous mutations at polymorphic sites. Together, this suggests that relapse-associated lineages likely undergo ongoing selection after the index infection and dissemination.

**Table 2:** Neutrality index calculations of the whole genome and bacteremia-associated genes for all MRSA bacteremia isolates and for relapse-associated lineages.

| Group | Fixed, Non-synonymous | Fixed, Synonymous | Polymorphic, Non-synonymous | Polymorphic, Synonymous | McDonald-Kreitman Value | P-value (G-test) |
| --- | --- | --- | --- | --- | --- | --- |
| All isolates - whole genome | 17271 | 24529 | 1381 | 1812 | 0.92 | **0.03** |
| All isolates - bacteremia genes | 232 | 330 | 19 | 22 | 0.81 | 0.53 |
| Relapse - whole genome | 2689 | 5326 | 45 | 12 | 0.133 | **< 0.01** |
| Relapse - bacteremia genes | 28 | 81 | 5 | 0 | — | — |

### Antibiotic resistance genotypes and phenotypes in relapses correspond with patient exposures

Eleven genes had unique SNPs with mutations in two or more relapse lineages (Fig. 4A). Mutations in these genes occurred regardless of CC, indicating that clade background alone did not contribute to mutations in these genes. Genes in which multiple relapse lineages showed non-synonymous mutations were implicated in known virulence traits and antibiotic resistance. The genes most commonly mutated were *mprF*, in five separate subject lineages, and *rpoB,* in four subject lineages. Changes in both *mprF* and *rpoB* are associated with drug resistance to antibiotics commonly used for bacteremia; we therefore investigated amino acid impact, isolate MIC, and treatment course. Multiple amino acid changes were detected between patients for proteins encoded by both genes. The same amino acid change, Ala477Asp, occurred for *rpoB* products in two subjects with rifampin resistance; for one subject the resistance phenotype was present in their first infection before relapse, a change at position 477 and position 471 (Asp471Tyr), which corresponded with a phenotypic change from susceptible to resistant (Fig. 4B). Three subjects with *mprF* mutations emerging in relapses demonstrated acquired daptomycin resistance with changes at Ser337Thr, Ser337Leu, and Leu291Ile (Fig. 4C). All recurrence-associated subjects with *mprF* mutations had exposure to daptomycin prior to mutation emergence regardless of daptomycin resistance phenotype. No other relapse lineages gained or lost resistance to rifampin or daptomycin without emergence of *mprF* or *rpoB* mutations.

**Figure 4:**
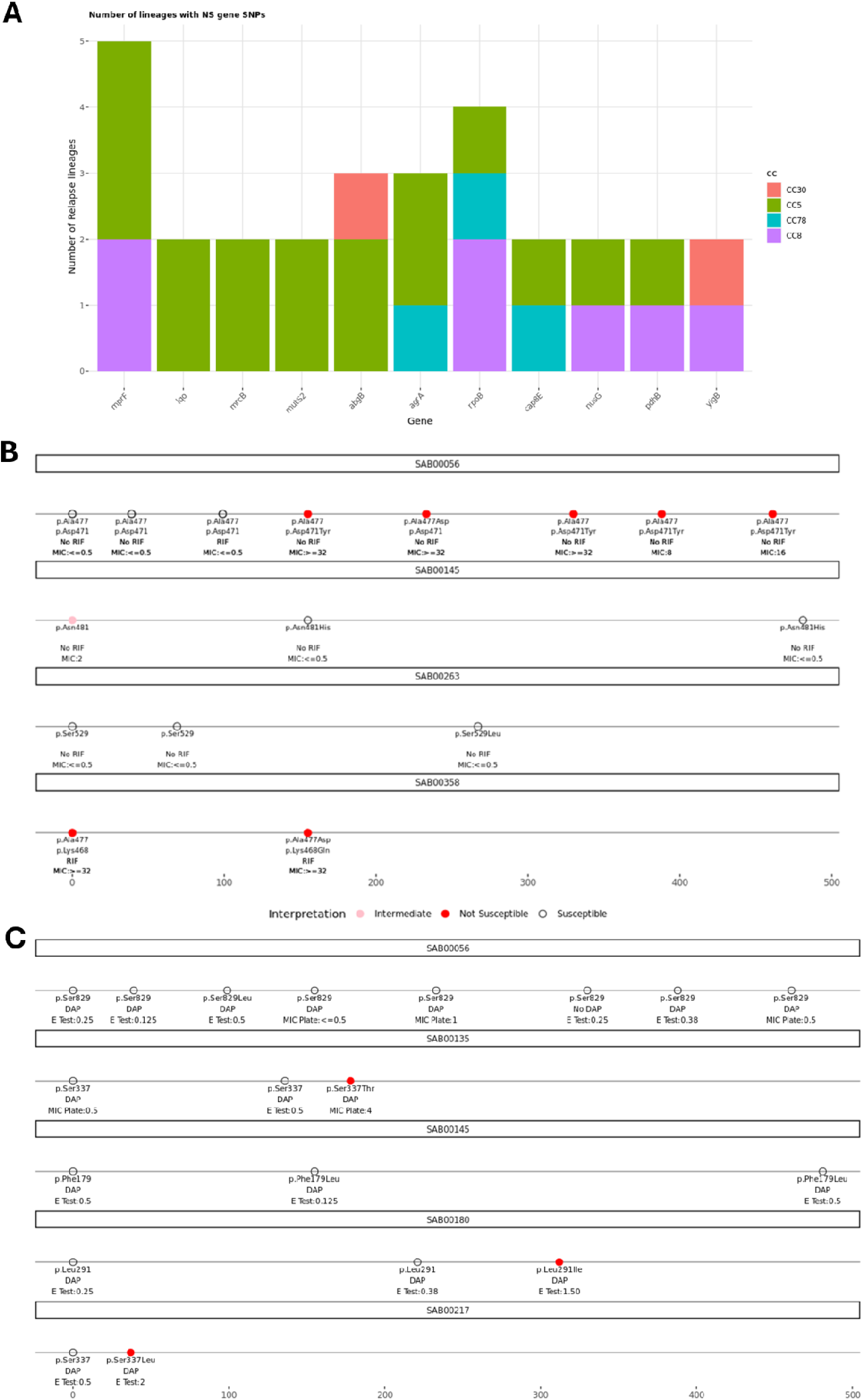
Commonly mutated genes among relapse lineages are associated with antibiotic resistance phenotypes. (A) Unique non-synonymous (NS) 354]mutations in coding regions of the genome were quantified by gene from isolates of relapse-associated infections and summarized relative to the subject from which the isolate was collected. Each unique mutation was annotated with the clonal background of the lineage from which the set of relapses were derived. For lineages with *rpoB* mutations (B) and *mprF* mutations (C), a timeline (days since index infection) was created for each set of relapsing infections by the subject experiencing that set of relapses. Individual episodes were annotated with the amino acid changes detected in the respective genes, the clinical assay used to assess minimum inhibitory concentration (MIC) and the corresponding MIC, and whether the patient was exposed to rifampin (RIF)(B) or daptomycin (DAP)(C). Dots are colored based on the clinical assay determination of drug susceptibility to RIF or DAP.

### Relapse isolates cluster with other subject isolates, but do not contribute to onward transmission

Nine clusters across subjects including at least one subject who had a relapse were identified. These clusters occurred in distinct lineages across CC5 and CC8 clades (Fig. 5). If relapse infections were contributing to onward transmission, or if patients were reinfected with a closely related circulating strain, we might expect that infections in unique hosts would cluster within relapse lineage clades. Across all nine clusters, isolates from different subjects clustered significantly outside of the relapse lineage isolates. This suggested that transmission occurred before the onset of relapsing infection and the unique lineages within a host were highly specific to the individual.

**Figure 5:**
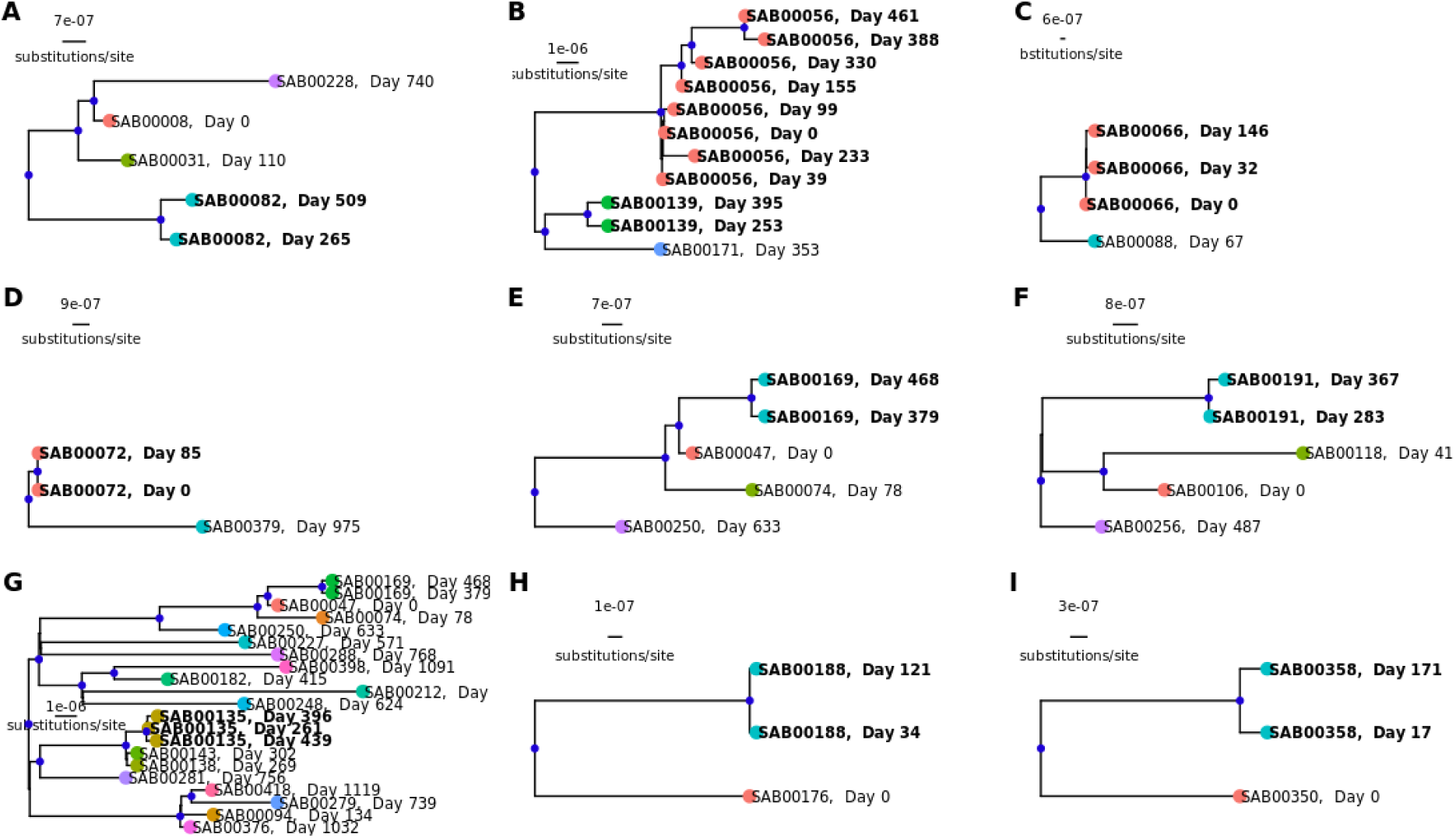
Non-relapse associated isolates cluster separately from relapse-associated isolates. A tree based on core genes was generated using the unequal transition/transversion rate plus empirical base frequencies model to investigate branching positions. Clusters were investigated when at least one relapse-associated isolate genome differed by 25 SNPs or fewer from an isolate from a different subject. Subtrees were extracted based on the most-recent common ancestor shared by relapse subject isolates and the clustered additional subjects. Nodes denoted with a blue dot indicate ultrafast bootstrap values and Shimodaira-Hasegawa approximate likelihood ratio test values that are greater than 70. Bolded tip labels indicate an isolate that is part of a relapse. Tips are annotated with the patient subject IDs and the number of days at which the isolate was collected relative to the earliest isolate in the cluster. Branch lengths are scaled in substitutions per site.

## Discussion

Our study showed that recurrent MRSAB is well differentiated into new and relapse infections by WGS and patterns of pathogen evolution. Consensus of a time and genomic definition of relapse is still not standard. It is imperative to document and compare genotypic and phenotypic patters among recurrent isolates that accompany different molecular definitions. For our investigation period shows that most isolates from the same person were separated by fewer than 25 core genome SNPs. We defined 3 additional isolates from episodes as relapses upon inspection of their phylogenetic relatedness to intrahost isolates. Previous studies have identified a similar pattern in distinguishing relapses from new infections even using less precise technologies including pulsed-field gel electrophoresis finding that approximately an equal number were classified as relapse and new infections [9]. Our SNP-based definition revealed a much larger percentage of relapse infections than previous studies would suggest. We observed unique phenotypic traits (i.e., daptomycin resistance) within relapse lineages, which would suggest emergence and likely persistence of a strain within the host. Though external reservoirs cannot be entirely ruled out, the nesting of the infections and the accumulation of genetic changes suggest a high likelihood of specific host-association. Furthermore, while phylogenetic histories of relapse-associated MRSAB isolates do occur in transmission clusters, they do not appear to be directly contributing to ongoing spread leading to additional MRSAB cases in healthcare settings.

Previous studies have identified risk factors for *S. aureus* bacteremia and MRSAB recurrence related to pathogen and host factors, and the type of care received. We identified similar risk factors in our dataset to those recorded previously, including a high prevalence of SCC*mec* type II [8] and a high proportion of unremoved foreign objects [6,7]. We did not identify a significant association between antibiotic treatment duration and recurrence. Other reports show a negative association between antibiotic duration and relapse [4,45]. These discrepancies may be explained by the type and duration of antibiotic treatment. Nevertheless, the number of recurrent infections we detected is from a single hospital group and may not be generalizable to other populations. Larger population sizes will be needed to identify variables with small effect sizes.

We found evidence that antibiotic therapy may have selected for strains with mutations in genes known to confer daptomycin and rifampin resistance. Several distinct amino acid changes were detected in isolates with both rifampin-resistant and daptomycin-resistant phenotypes at the index infection and acquired in some, as demonstrated in relapse isolate genomes. Changes to the amino acid sequence corresponded with expected phenotypic changes observed in other studies such as the changed asparagine at position 481 of the *rpoB* transcription product resulting in a rifampin-intermediate resistance profile [47], and the point mutations observed in *mprF* that resulted in changes at the same amino acid site (Ser337Thr and Ser337Leu) corresponding to daptomycin resistance. Multiple amino acid changes corresponding with phenotypic changes here demonstrate wide diversity of mutational profiles of *rpoB* and *mprF* even within a single study population and convergence of traits within infected patients rather than single, resistant strains spreading between individuals.

Management of MRSAB involves a comprehensive assessment of patient history, physical examination, and source identification [1]. Delays removing or draining an infection focus can increase the risk of persistent bacteremia [1] or metastatic spread [48]. When central venous catheters or other foreign bodies are suspected to be the source of SAB, removal of the foreign body is considered, though it is not always possible when it increases risk of patient harm. We found genomically similar MRSAB episodes common among patients with foreign body infections, suggesting the presence of persistent infection foci reseeding the blood. We had previously reported MRSA infections associated with a foreign body have a nearly 5-fold greater odds among patients who had a previous MRSA infection within a year compared to those with no previously reported infection [49]. Clinicians should maintain a high index of suspicion of implanted foreign bodies as a source of recurrent MRSAB and advise patients accordingly during MRSAB recovery.

WGS provided additional support for case-identification of new and relapsing infections in the context of the clinical history. The complexity of host factors, within-host selection pressures, strain background and type of antibiotic treatment all play interacting roles in relapse emergence. It is not be possible to determine if recurrent MRSAB is caused by the same strain without performing WGS. Examining isolates in a population context further differentiated relapses from new infections. Relapsing isolates undergo positive adaptation to the host, and convergently mutating genes are consistent with long-term usage or high exposure to antibiotics, but other traits necessary for survival in the body may still play an important role in persistence. Additional work is necessary to understand survival duration of *S. aureus* strains across individual hosts’ body sites. Combining the frequency of genetic mutations, genetic relatedness, and known clinical risk factors for recurrence will lay the groundwork for better prediction of new MRSAB and other *S. aureus* bacteremia relapses and may contribute to therapy strategies to prevent relapsing infections.

## Data Availability

All data produced are available online at https://github.com/bmtalbot/bacteremia_recurrence

https://doi.org/10.5281/zenodo.15066414

## Acknowledgements

The authors would like to thank Pam Tolomeo, MPH, of the University of Pennsylvania for assistance with clinical data management and The Penn/Children’s Hospital of Philadelphia (CHOP) Microbiome Center for whole genome sequencing services.

## Author contributions

BMT: Conceptualized project, designed analysis, collected data, analyzed data, drafted initial manuscript, reviewed and approved manuscript.

NFJ: collected data, reviewed and approved manuscript.

KH: collected data, reviewed analysis, reviewed and approved manuscript.

TA: Collected data, reviewed and approved manuscript.

KB: Collected data, reviewed and approved manuscript.

MZD: Conceptualized project, designed analysis, collected data, drafted the initial manuscript, reviewed and approved manuscript.

TDR: Conceptualized project, designed analysis, drafted the initial manuscript, reviewed and approved manuscript.

## Data availability

Raw sequence reads for this study are publicly available in the Sequence Read Archive under the project ID PRJNA751847. Metadata and analysis code used to generate the figures and tables are available at https://doi.org/10.5281/zenodo.15066414

## Funding

TDR and MZD were funding by NIAID awards NIH 1R01AI139188-01 (NIAID) and NIH 1 R01 AI158452-01A1. BMT and TDR were supported by the Office of Advanced Molecular Detection, Centers for Disease Control and Prevention Cooperative Agreement Number CK22-2204 through contract 40500-050-23234506 from the Georgia Department of Public Health. The funders had no role in study design, data collection and analysis.

## Conflicts of Interest

The authors report no conflicts of interest.

## Notes

### Competing Interest Statement

The authors have declared no competing interest.

### Author Declarations

This study was considered exempt by the University of Pennsylvania Institutional Review Board.

