## Supplemental Figures and Tables for "Genomic investigation of MRSA bacteremia relapse reveals diverse genomic profiles but convergence in bacteremia-associated genes"

**Supplemental Material**

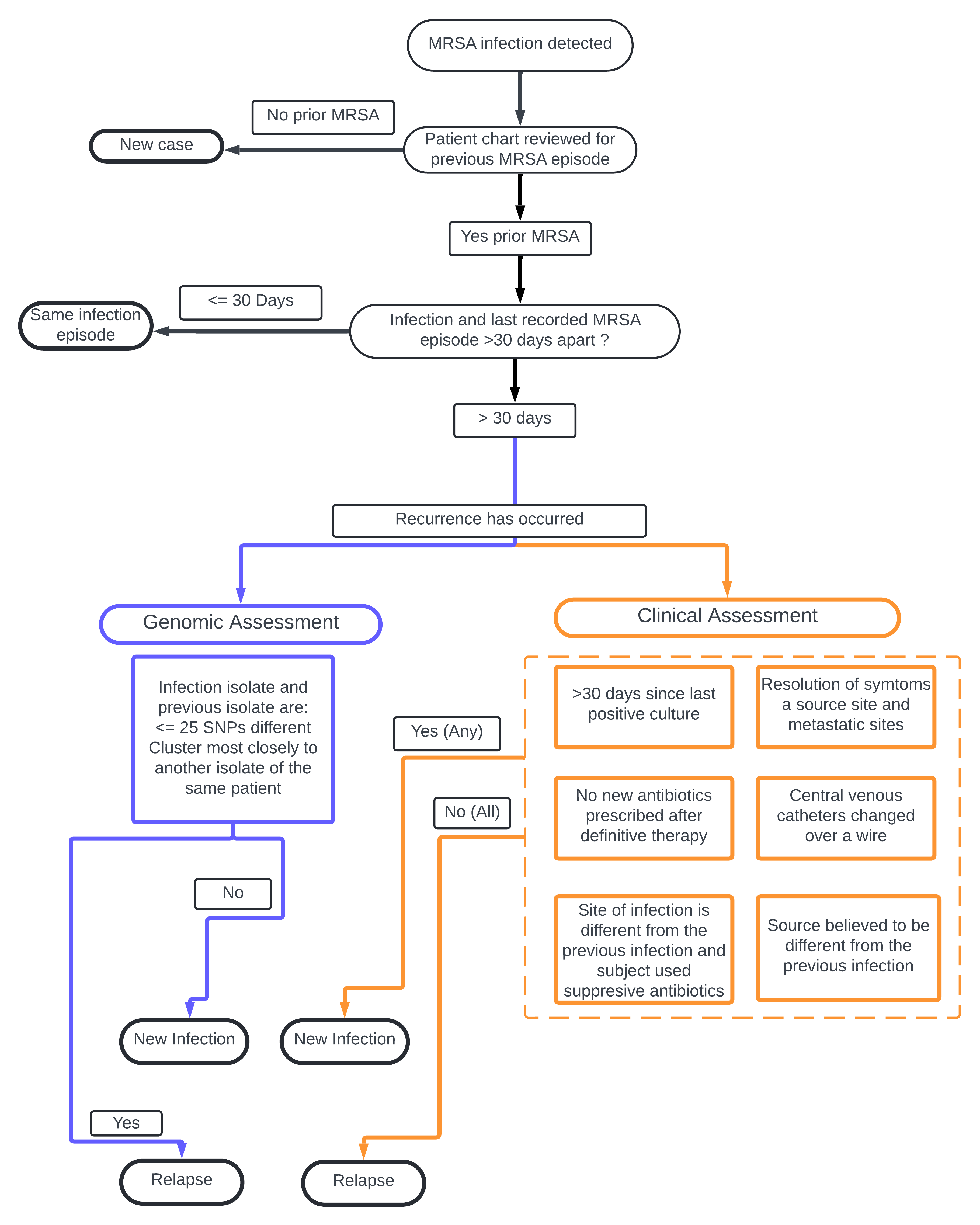

**Figure S1: Clinical and Genomic Criteria for defining Recurrent new and relapsing infections.** The flowchart is formatted as a decision tree with round ovals indicating terminals for branch points and rectangles indicating options that correspond with the most recent terminal. Black lines indicate criteria assessed regardless of the definition. Blue lines indicate criteria assessed for genomic definitions, and orange lines indicate criteria that fulfill the clinical definitions.

**Table S1. Clinical and demographic characteristics at the time of the first MRSA bacteremia episode among survivors versus decedents during their index episode**

| Patient Attribute | Overall (N = 397) | Survived (N = 311) | Died (N = 86) | P-Value |
| --- | --- | --- | --- | --- |
| **Age at Diagnosis (median, IQR)** | 56 (43-68) | 54 (41-65) | 67 (52-75) | <0.001^a^ |
| **Patient Demographics** | | | | |
| **Sex** |  |  |  | 0.1388 ^b^ |
| Male | 224 (56%) | 182 (58%) | 42 (49%) |  |
| Female | 173 (44%) | 129 (41%) | 44 (51%) |  |
| **Race** |  |  |  | 0.7775^c^ |
| *White* | 186 (47%) | 146 (47%) | 40 (47%) |  |
| *Asian* | 8 (2%) | 5 (1.6%) | 3 (3.5%) |  |
| *Black* | 165 (42%) | 131 (42%) | 34 (40%) |  |
| *More than one race** | 3 (0.8%) | 2 (0.6%) | 1 (1%) |  |
| *Other Race* | 12 (3%) | 9 (3%) | 3 (3.5%) |  |
| *Don’t know/refused* | 23 (6%) | 18 (6%) | 5 (6%) |  |
| **Ethnicity** |  |  |  | 1^c^ |
| *Hispanic or Latino* | 17 (4%) | 14 (4.5%) | 3 (3.5%) |  |
| *Not Hispanic or Latino* | 380 (96%) | 297 (95.5%) | 83 (96.5%) |  |
| **Patient Comorbid Conditions** | | | | |
| **Chronic skin disease** | 36 (9%) | 32 (10%) | 4 (5%) | 0.1372^c^ |
| **Diabetes** | 137 (34.5%) | 105 (34%) | 32 (37%) | 0.6405^b^ |
| **Cancer** | 65 (16%) | 25 (8%) | 40 (46.5%) | 0.0006022^b^ |
| **Respiratory disease** | 93 (23%) | 68 (22%) | 25 (29%) | 0.2104^b^ |
| **Cardiovascular disease** | 195 (49%) | 144 (46%) | 51 (59%) | 0.04416^b^ |
| **Infective endocarditis** | 66 (17%) | 50 (16%) | 16 (19%) | 0.7502^b^ |
| **Liver disease** | 40 (10%) | 30 (10%) | 10 (12%) | 0.1385^b^ |
| **Kidney disease** | 111 (28%) | 83 (27%) | 28 (32.5%) | 0.3483^b^ |
| **Hemodialysis in the last 12 months?** | 75 (19%) | 59 (19%) | 16 (19%) | 1 |
| **Current intravenous drug use** | 81 (20%) | 74 (24%) | 7 (8%) | 0.0008494^c^ |
| **Complication of Bacteremia** | | | | |
| **Infective endocarditis** | 66 (17%) | 50 (16%) | 16 (19%) | 0.7502^b^ |
| **Epidural abscess** | 21 (5%) | 18 (6%) | 3 (3.5%) | 0.6765^c^ |
| **Soft tissue abscess** | 46 (12%)) | 41 (13%) | 5 (6%) | 0.1009^c^ |
| **Osteomyelitis** | 77 (19%) | 67 (21.5%) | 10 (12%) | 0.0817^c^ |
| **Septic pulmonary emboli** | 50 (12.5%) | 41 (13%) | 9 (10%) | 0.6745^c^ |
| **Other septic or metastatic complication(s)** | 53 (13%) | 43 (14%) | 10 (12%) | 0.7209^c^ |
| **Involvement of foreign body in the bacteremia** | 84 (21%) | 69 (22%) | 15 (17%) | 0.4212^b^ |
| **Healthcare Acquisition of index** |  |  |  | 0.01158^b^ |
| *Community Associated* | 46 (12%) | 38 (12%) | 8 (9%) |  |
| *Healthcare Associated* | 91 (23%) | 61 (20%) | 30 (35%) |  |
| *Healthcare Associated- Community Onset* | 260 (65.5%) | 212 (68%) | 48 (56%) |  |
| **How many antibiotics was the patient exposed to during their index infection?** | 1 (1-2) | 2 (1-2) | 1 (1-2) | 0.01342 |
| Vancomycin | 373 (94%) | 295 (95%) | 78 (91%) | 0.1974^c^ |
| Daptomycin | 148 (37%) | 123 (40%) | 25 (29%) | 0.07^c^ |

a Kruskal-Wallis test

b Chi-square

c Fishers Exact

**Table S2: List of *S. aureus* Bacteremia-associated genes and from peer-reviewed literature**

| **Gene** | **Product** | **Isolate Source** | **Allelic or mutational change detected** | **Phenotype of mutation** | **Literature Support** | **Literature refute/no evidence** |
| --- | --- | --- | --- | --- | --- | --- |
| ACME (arc and opp3) | arginine catabolic mobile element | clinical isolates |  | Presence increased pathogenicity in bacteremia model (rabbits) | Diep 2008a [1] |  |
| Agr | virulence regulator | clinical isolates |  | Increased genetic diversification compared to agr+ strains and colonizers; dysfunction leads to increased VAN MICs | Altman 2018b[2],  Cheung 1994 [3],  Tsuji 2009 [4],  Chong 2013 [5] |  |
| AgrA |  | clinical isolates | Non-synonymous, truncation | High-level rifampin resistance | Hachani 2023 [6],  Giulieri  2018 [7],  Benoit 2018 [8] | Howden 2008, [9] |
| AraC | AraC family transcriptional regulator | clinical isolates | premature stop | Untested, but present in the isolates not from nasal carriage | Young 2012 [10] |  |
| ausA |  | clinical isolates | Non-synonymous, truncation | Escape from epithelial cell endosomes | Hachani 2023 [6] |  |
| clpX |  | clinical isolate | Non-synonymous | Reduction of expression of virulence | Baek 2015 [11] |  |
| cna | collagen-binding adhesin | clinical isolates | non-synonymous | Decreased attachment to collagen | Iwata 2020 [12] |  |
| coa | coagulase | laboratory strain | deletion | Loss of coagulase function, decreased virulence | Liu 2021 [13],  Altman 2018 [2] |  |
| dfrB |  | Clinical isolates | Non-synonymous | Trimethoprim resistance | Young 2021 [14] |  |
| edinB | epidermal cell differentiation inhibitor | laboratory strains |  | Presence of gene increases ADP-ribosylation, increased prevalence of bacteremia during pneumonia and bacterial load | Courjon 2015 [15] |  |
| ess | virulence regulator | clinical | insertion sequence | Increased expression of ESAT^-like secretion system virulence factors (when agr-) | Altman 2018 [2] |  |
| essB | ESAT-6 secretion system component | laboratory strains of ST398 |  | Deletion results in decreased neutrophil killing and lethality in blood | Wang 2016 [16] |  |
| fnbA | fibronectin-binding protein A | clinical isolates | Non-synonymous SNPs | Enhanced binding to fibronectin, associated with cardiac device infections from bacteremia isolates [[62]](https://www.zotero.org/google-docs/?ehk9EH) | Hos 2015 [17] |  |
| fusA |  | clinical isolates |  | Fusidic acid resistance by target alteration, some small evidence that one isolate arose from an SCV phenotype | Lannergard 2009 [18] |  |
| fusB |  | clinical isolates |  | Fusidic acid resistance by protecting translation apparatus | Lannergard 2009 [18] |  |
| fusC |  | clinical isolates |  | Fusidic acid resistance by protecting translation apparatus | Lannergard 2009 [18] |  |
| hglABC |  | laboratory strians |  | Presence assists with survival in blood | Malachowa 2011 [19] |  |
| ica | intracellular adhesin locus conferring poly-N-acetylglucosamine production | laboratory strains |  | Loss of function shows a greater susceptibility to Ab-dependent killing by leukocytes | Kropec 2005 [20] |  |
| katA | catalase enzyme | clinical isolate | stop codon and truncation | Loss of catalase activity but still results in septic arthritis | Lagos 2016 [21] |  |
| lukED | leucotoxin ED | laboratory strains |  | Targets murine phagocytes leading to cytotoxic effects at infection site | Alonzo 2011 [22] |  |
| mgrA | virulence regulator | laboratory strains |  | Association of loss of function leads to increased susceptibility to host defense response cells via *mprF* and *dltA* expression, possibly loss of *fnbA* expression, and general decreased impact on host health in a bacteremia model; mutation leads to increased virulence in a mouse model | Li 2019 [23],  Rom 2017 [24] | Howden 2008 [9] |
| mprF | cell membrane structure | clinical isolates | non-synonymous, deletions, | Increased daptomycin resistance | Ji 2020 [25],  Baek 2015 [11],  Chen 2015 [26] |  |
| mpsB | cation translocation in cell membrane | clinical isolates; laboratory strains |  | Small colony phenotype, suppression of agr activation due to lowered membrane potential | Douglas 2021 [27] |  |
| mspA | membrane protein | clinical and laboratory |  | Role in toxin production, resistance to innate immue cells, and iron homeostasis | Duggan 2020 [28] |  |
| parC | topoisomerase IV | clinical isolate | insertion | confers quinolone resistance (e.g., CIP) | Gao 2010 [29] |  |
| psm-mec | phenol-soluble modulin alpha type | clinical isolates | promoter SNP | Decreased biofilm formation and increased PMSa3 and Hld expression | Aoyagi 2014 [30] |  |
| purR | purine biosynthesis regulation, and regulation of fibronectin binding protein | laboratory strain | Non-synonymous SNP | Increased clumping in blood related to fibronectin binding, increases in SarA expression which has other virulence factor down stream events | Goncheva 2019 [31],  Alkam 2021 [32] |  |
| PVL (lukS/F) | Panton-Valentine leukocidin | laboratory strains |  | Increased pathogenesis in early stages of bacteremia | Diep 2008ab [33] |  |
| rel | synthesis of (p)ppGpp during AA starvation | clinical isolates | Four non-synonymous (D134Y, A301T, E384K, V670G), and a 4-bp deletion encompassing codon N697 that results in a frameshift causing a premature stop codon at position 701; Gao showed a Phe 128 Tyr substitution from a nucleotide substitution) | Shortened lag phase, increased fitness in nutrient-poor conditions; increased resistance to antimicrobials and defensins; possibly related to agr upregulation for Gao 2010 | Chen 2023 [34],  Bryson 2020 [35],  Gao 2013 [36],  Gao 2010 [29] |  |
| rlmN | ribosomal RNA large subunit | clinical isolate | insertion | Confers linezolid resistance, uniquely from previous 23 rRNA mutations | Gao 2010 [29] |  |
| rot | regulation | laboratory strain |  | Change in sepsis virulence (i.e., survival) in mice, background dependent | Rom 2021 [37],  Rom 2017 [24] | Howden 2008 [9] |
| RpiRc |  | laboratory strain (USA300-LAC) | altering protein expression | Repressed RNAIII to mimick rot deletion, leading to bloodstream infection phenotype | Balasubramanian 2016 [38] |  |
| rpoB | RNA polymerase |  | Non-synonymous |  | Giulieri 2018 [7],  Baek 2015 [11],  Gao 2013 [36],  Gao 2010 [29],  Villar 2011 [39] |  |
| rpoD (sigA) | RNA polymerase sigma factor | laboratory strain | excision of an IS256 element | Decreased capacity to infect bone and increased virulence in mouse | Suligoy 2020 [40] |  |
| rsp | transcription factor repressor of surface protein | clinical isolates | Non-synonymous and premature stop codon | Reduced lethality, reduced cytotoxicity | Das 2016 [41] |  |
| SaeRS | regulatory system | laboratory strain | deletion | Increased survival in mouse blood; in balance with sarA protease production for virulence; in lab, lack of SaeRS increases mortality, but down regulation of leukocidins and immunomodulatory genes, and some adhesion genes) | Liu 2021 [13],  Beenken 2014 [42],  Nygaard 2010 [43] | Voyich 2009 [44] |
| sar | virulence regulation | laboratory strain |  | Loss of virulence factors leading to loss of attachment to heart valves; loss of virulence that may be protease-mediated | Cheung 1994 [3],  Beenken 2014 [42],  Zielinska 2012 [45] |  |
| SCC*mec* type IV | MGE harboring *mecA* | clinical isolates |  | Higher association with bacteremia or central line infections | Nakano 2022 [46],  Young 2021 [14] |  |
| selw | production of SAg SelW | clinical isolates; laboratory strains |  | Superantigen activation of T-cell proliferation | Vrieling 2020 [47] |  |
| srtA | sortase A | laboratory strains |  | Deletion results in reduced mortality and dissemination to tissues after introduction to the bloodstream in an injection introduction | Wang 2015 [48] |  |
| stp |  |  |  |  | Giulieri 2018 [7] |  |
| thyA | thymidylate synthase gene needing THF cofactor | clinical isolate | premature stop | Small colony phenotype, related to adaptation of trimethoprim-sulfamethoxazole resistance? | de Souza, 2020 [49] |  |
| xerC | recombinase | laboratory strain (USA300-LAC and UAMS-1) |  | Mutation results in decreased bacterial load in murine bacteremia, decreased accumulation of alpha toxin, and decreased biofilm production | Atwood 2016) [50] |  |
| yycH | modulation/downstream of walK which is important for cell wall metabolism | clinical isolates | frameshift, premature termination | VAN and DAP nonsusceptibility, mechanism unknown or multifaceted | Chen 2015 [26] |  |
|  | Wall teichoic acids | laboratory strain |  | Loss of function results in less adherence to endothelial cells and proliferation to spleen and kidneys | Weidenmaier 2005) [51] |  |
